# Visual Acuity alterations in heavily impaired Congenital Zika Syndrome (CZS) children

**DOI:** 10.1101/2022.02.09.22270174

**Authors:** Luiz C. P. Baran, Diego da S. Lima, Leonardo A. Silva, Heydi S. Tabares, Sarah L. Dias, Andrea Araújo Zin, Maria Elisabeth Lopes Moreira, Marcelo F. Costa, Dora F. Ventura

## Abstract

We aim to assess visual acuity (VA) in Congenital Zika Syndrome (CZS)-children to evaluate visual loss. To that end we evaluated 41 CZS- children, from Rio de Janeiro using Teller Acuity Cards. They had Zika virus-infection confirmed by reverse transcription–polymerase chain reaction (RT-PCR) or clinical evaluation. VA below normative values was present in 39/41 (95%). In 10 cases, VA was only marginally below normal; in the remaining 29 cases, VA was more than 0.15 logMAR below the lower limit. There was no relationship between VA and cognitive domain tasks, although there was a relationship between VA and motor domain tasks. Thirty-seven children performed at least one task in the cognitive set, 14 children did not perform any task in the motor set. Children with VA above the lower limit performed better in the cognitive and motor tasks. We concluded that ZIKV- infected children with CSZ were highly VA impaired which correlated with motor performance, but not with cognitive performance. Part of the children had VA within the normal limits and displayed better performance in the cognitive and motor set. Therefore, even if heavily impaired, most children had some degree of visual acuity and visual function and may benefit from visual rehabilitation.

## 1 Introduction

Gestational Zika virus (ZIKV) infection may lead to Congenital Zika Syndrome (CZS)^1–4^, with microcephaly as its most known manifestation. In Brazil, where the pandemic had a high impact, a great number of cases of microcephaly were reported in 2015-2016 ^5^. There was a clear asymmetry in regional occurrence of CZS, which was more frequent in the Northeast region of Brazil, and also in Rio de Janeiro and Cuiabá. A causal link between the ZIKV infection and the occurrence of microcephaly was inferred by temporal correlation and was later experimentally confirmed^1,6,7^. CZS, is not restricted to microcephaly and has a myriad of manifestations, due, chiefly to neurological impairment and massive intracranial volume loss^1^. Features of the CZS spectrum include partially collapsed cranium, neurological effects such as thin cerebral cortices, seizures, polymicrogyria and subcortical calcifications, increase in cerebral fluid spaces (ventriculomegaly), hypoplasia or loss of the corpus callosum, decreased myelination, cerebellar hypoplasia and brainstem and basal ganglia calcifications, along with somatic abnormalities such as hypertonia, limb contracture, arthrogryposis (joint stiffening), altered craniofacial proportions, spasms, irritability, problems in swallowing and hearing losses^1,3,4^.

CZS also affects the visual system. Clinical manifestations include chorioretinal atrophy, macular pigmentary mottling, vascular changes, retinal focal spots, optic nerve anomalies, microphthalmia, iris coloboma, cataracts and intraocular calcifications^1,2,8–10^. Infection of central nervous system might incur in eye motility issues such as strabismus, nystagmus and accommodative capacity impairment as well^11^.

Even though some studies focusing on visual and ocular alterations due to ZIKV-infection have examined the impact of these vision threatening events since the beginning of the recent ZIKV epidemics^2,8,10,12–18^, the full scope of CZS effects on the visual system is not completely characterized yet. In a cohort in the Northeast of Brazil, where the highest rate of children born with CZS during the 2015-2016 ZIKV epidemy was seen. Ventura and collaborators^2^ reported visual acuity losses in 76% of children with CZS (n=25; all with microcephaly); failure in the detection of a low-contrast pattern in 65% (n=31) and failure to achieve at least one visual development milestone in 97% (N=31) of tested children, apart from eye movement conditions. The authors found VA losses in all 11 tested children, with acuity values ranging from 0.5-5 octaves below lower limits. Ventura et al.^12^, found VA deficits in approximately 85% of a larger sample (N=119). Those studies were carried out with populations from North-Eastern Brazil.

The impact of CZS on VA was also assessed, by our team, in a cohort from South-Eastern Brazil (Jundiaí, São Paulo), a region that presented a different epidemiological profile: although ZIKV infection rates in the general population were high, the incidence of CZS was relatively low^19^. Baran et. al^17^ found that babies exposed to maternal ZIKV infection during pregnancy that had not become infected had VA within normal limits. Conversely, in the group of babies that acquired infection during gestation, 5/24 children (21%) had VA impairment, 2 of which with microcephaly. Moreover, examined as group, the infected children showed a slower VA development rate compared with the control and exposed groups^17,18^.

Visual deficits were also observed in a sample from the state of Rio de Janeiro^10,14,15^, the region with the highest incidence of CZS outside North-Eastern Brazil^19^. In this sample, 30% of the patients (N=173) did not meet the requirements of a visual screening test that assessed the child’s capacity to fixate a single monochromatic pattern and follow it with the gaze (The Fix-and-Follow test). This exam, while useful for identifying children with severe VA deficits, does not yield a threshold estimate, required for a direct comparison with previous studies. Moreover, since the pattern used in the test has low spatial frequency, mild VA losses might go undetected.

In the present study, we measured VA in infants and children who had been exposed to ZIKV during gestation and developed CZS. Our aim was to better characterize the incidence and magnitude of vision loss in CZS patients by assessing VA - a quantitative and universally measured indicator of visual function. VA was assessed behaviorally using a clinical version of Teller Acuity Card (TAC)^20–25^. The TAC procedure is well-established as an efficient and reliable instrument to measure VA in young children in a clinical setting^21–24^.

Here we add to our previous works^17,18^ and to the literature of the area by evaluating patients that were at an older age (previous studies had patients ranging from 4-13 months of age ^2,11,12,^ while patients in the present study had a median age of 24 months); more severely affected by CZS; by characterizing VA function in affected children from a different geographic region not covered by previous work, with different epidemiological profile and enrolled in a cohort which followed a different study design.

By examining a population from another region, selected with a different selection criteria, in a cohort of older, more impaired children, we further characterize the spread of the virus in Brazil, continuing to help document the diversity of the ZIKV’s impact in different regions of the country. These differences may be due to a variety of factors, such as strain differences, host susceptibility ^26^, water contamination^27^, or malnutrition^28^. Additionally, it is critical to document and characterize the visual function alterations in CZS children with and without microcephaly.

## 2 Materials and Methods

This research is in line with the tenets of the Declaration of Helsinki^29^ and was approved by the Ethics Committee for Human Research of the University of São Paulo’s Institute of Psychology (number 67031216.0.0000.5561) and by the Ethics Committee for Human Research of the Institute Fernandes Figueira (IFF) – Fiocruz (number 526756616000005269). An informed consent was signed by the parent or accompanying adult of the child after being given an explanation on the nature and purpose of the study.

Children examined were part of the Vertical Exposure to Zika Virus and Its Consequences for Child Neurodevelopment cohort, registered under NCT03255369 at the NIH Clinical Trials Database^14,15,30^. Methodological details about the enrollment criteria for patients can be found at the Clinical Trials page for the cohort (https://clinicaltrials.gov/ct2/show/NCT03255369).

The cohort was assembled and followed clinically in the Institute Fernades Figueira (IFF) Fiocruz, by the Institute’s research team under the guidance of coauthor AZ. For the present study we recruited children born from mothers with suspected ZIKV-infection during pregnancy (such as rash, arthralgia, myalgia and fever) and who filled one or more of the following criteria: (a) Positive RT-PCR sample for ZIKV, from either pregnant mother or from the child within 10 days after birth; (b) Presence of structural congenital alterations detected via ultrasound; (c) Clinical manifestations typical of CZS (such as microcephaly and eye alterations) detected via clinical examination after birth. The RT-qPCR, serologic test and clinical examination (including fundoscopic evaluation) for ZIKV infection was performed by IFF Fiocruz research team^14,15,30^.

The TAC test consists of 15 gray 25.5 × 55.5 cm cards (35% reflectance). Each card has a small peephole (4 mm in diameter) in the center to allow the experimenter to observe the child’s looking behavior. Each TAC card contains a 12 × 12 cm square-wave grating (black and white stripes, at approximately 95% contrast) on one side of the central peephole. Gratings range from 0.32 cycles/cm to 38.0 cycles/cm in approximately half-octave steps. The space-averaged luminance of each grating is equal to the card’s gray background.

Patients sat in an adult’s lap facing an Observer holding the card. The cards were presented by the Observer to the child from a distance of 38 cm. All children were examined at the distance of the 38 cm, despite their age because of the attentional, neurological and eye motility issues (such as nystagmus) of the children evaluated in this study. At the start of each acuity measurement, the Observer was always blind to the left-right location of the grating on the card. The Observer attracted the child’s attention to the card and watched the direction of her gaze through the peephole. The Observer ‘s task on each trial was to make a forced-choice guess about the location of the grating based on the child’s behavior (primarily the direction of gaze). An Assistant, behind the child, who could see the grating’s location, recorded the Observer ‘s guess on each trial and gave him feedback.

A 1-Down, 1-Up staircase procedure was used during testing. First the card with the 0.23 cy/cm spatial frequency stymulus (first card in the set) was presented to account for the possibility that children with very low acuity could be in the sample. A card with half-octave higher spatial frequency was selected every time the Observer made a correct guess about the grating position, and a card with a half-octave lower spatial frequency every time the Observer made an incorrect guess. The staircase was completed after a minimum of three reversals depending on the experimenter’s confidence about his judgment of the children’s responses. VA threshold was calculated as the geometric mean of the spatial frequencies of the gratings in the final 3 reversals.

VA thresholds were converted to logMAR based on the distance of 38 cm common across all participants. If participants had a prescription for refractive correction, and used well adapted spectacles, they performed the test wearing them. Children born prematurely had their age corrected (from post-natal to post-term), and their VA was compared to post-term age norms assuming that no differences in VA between terms and preterm^31–35^.

Children were subjected to two sets of brief tasks of functional vision assessment, fifteen items related to the visual function of the Bayley Scales of Infant and Toddler Development, Third Edition (Bayley-III, 2006) were selected, nine in the cognitive set and six in the motor set, in the same occasion as their VA was being tested, with the aim of examining if there was any relationship with the measured VA. A complete application of the developmental outcomes for the patients in the cohort (at a younger age) has been published previously^36^.

VA values were compared with normative values established by Salomão & Ventura^22^, using the same tolerance intervals. The patients were categorized accordingly to their visual acuity falling above, marginally below and below normative values. Effects of both the patient’s age and the presence of any kind of retinal damage in VA was investigated by estimating the coefficients of a linear regression model. Confidence intervals for the coefficients and the Likelihood Ratio (LR) Statistic^37^ were calculated. The relationship between acuity and functional visual outcomes was evaluated via a logistic regression statistical model. We established the criteria of having successfully completed at least 2/3 of the investigated visual acuity tasks as the binary outcome for the model which was related to the VA value (in logMAR) as the independent variable. Any associations were considered significant if the calculated LR statistic value was above the 0.05 significance level. The statistical routines from the statsmodels library^38^ were used for all statistical procedures.

## 3 Results

We evaluated 45 children, from which 41 met our inclusion criteria (20 boys). The age range was 21-34 months. Diagnostic of CZS based on a positive RT-PCR test result from either the mother and/or the child was available in 18 cases. For the remaining 23 cases, diagnosis was based on the pregnant mother having ZIKV symptoms and the child presenting CZS outcomes, such as microcephaly. Within the group of children clinically diagnosed with ZIKV-infection but without positive laboratory confirmation, either the child or the mother had negative results in 14 cases at the time of testing (which does not rule out viremia at an earlier stage relative to the testing occasion), while 9 cases were untested. There were 26 co-occurrences of microcephaly and retinal damage, and a single case of ZIKV-infected child with eye retinal damage in the absence of microcephaly. In all cases included based on clinical criteria, the mothers tested negative for the exposure to TORCH agents.

The patients in the sample had a mean visual acuity of 1.0 logMAR (SD = 0.3 logMAR; Range = 1.73 to 0.5 logMAR). Most children (39/41–95%) had VA below normative values for their age. In 10 cases, VA was only marginally below normative values (visual acuity within 0.15 logMAR from the lower normative limit, equivalent to a single spatial frequency step in the card set); for the remaining 29 cases, visual acuity was more than 0.15 logMAR below the lower limit (Figure 1).

**Figure 1.**
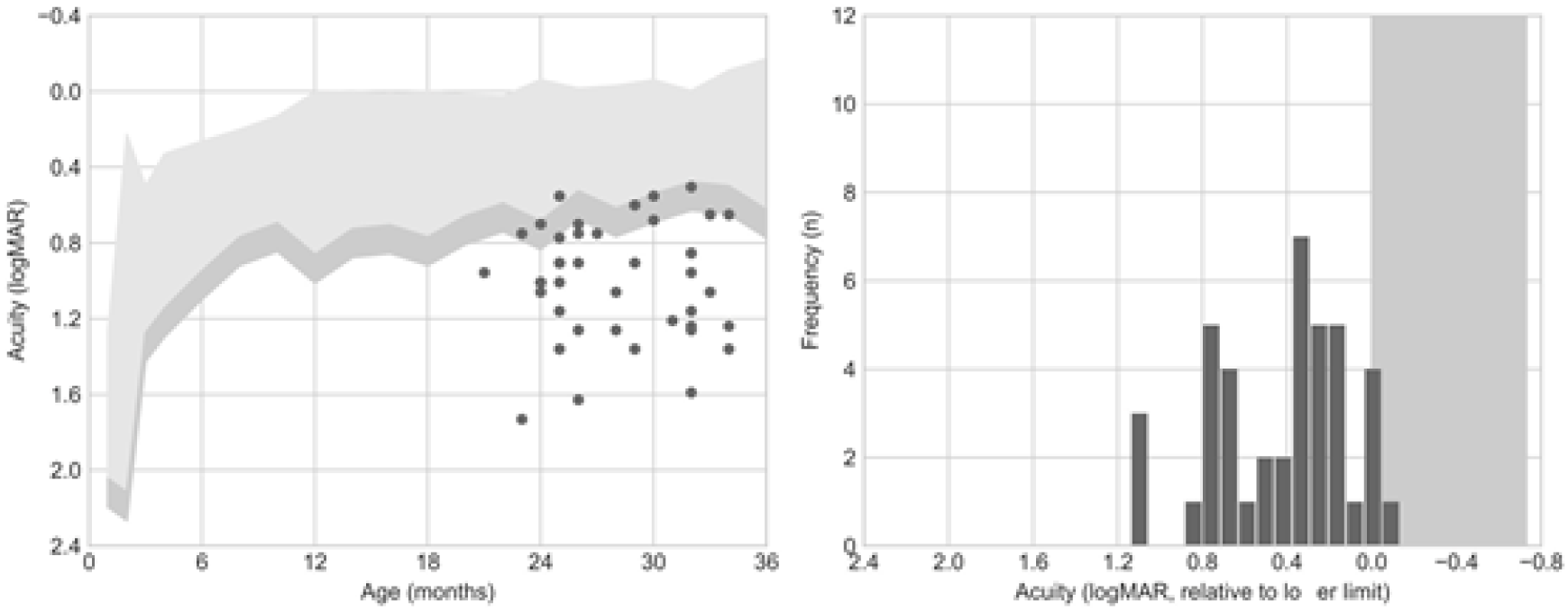
Visual acuity (VA) outcomes. The left panel shows VA compared to the normative values published by Salomão & Ventura (1995). The right panel shows acuity as an offset from the mean acuity for the patient’s age. 6/41 (15%) of patients presented an acuity value falling below this criteria.

VA measurements within 0.15 logMAR of the lower normative limits require repeated testing and/or additional clinical information collected via other techniques before diagnosing VA deficits (Stereo-Optical Co., 2005). Among the ten children with VA only marginally below normative values, seven (70%) presented retinal damage. In the group of children with VA more than 0.15 logMAR below inferior limits, 21 (72%) presented retinal damage. The mean and range of VA values, and their relation to retinal damage, using this classification is summarized at Table 1.

**Table 1.** Visual acuity (VA) by classification.

| Classification | N | Minimum Acuity<br>(logMAR)* | Maximum Acuity<br>(logMAR)* | Eye<br>damage (N) |
| --- | --- | --- | --- | --- |
| Above inferior limit | 2 | 0.6006 | 0.5527 | 1 |
| Marginally below inferior limit (< 0.15 logMAR) | 10 | 0.9574 | 0.5048 | 7 |
| Below inferior limit (> 0.15 logMAR) | 29 | 1.7316 | 0.7010 | 23 |

A multiple linear regression model that included age (in months) and presence of any retinal damage as independent variables to predict VA (in logMAR) showed no significant effect for either variable (Age = −0.0278 to 0.0237 logMAR/month; Retinal Damage = −0.0564 to 0.03424 logMAR change; LR = 2.08; p =0.354). In the age range tested (21-34 months) only a very modest increase in visual acuity is expected (0.2 logMAR in the average of the normative values), consequently, it is not possible to evaluate the problems in VA development in this sample. Since all children were tested binocularly, the absence of a relationship between eye damage and acuity could be due to compensation by an unaffected eye.

Functional vision examination results are summarized in Table 2. Overall, most patients were able to complete simpler tasks (pay attention to object and react to the examiner’s face occlusion) but only a few completed more complex tasks (persistent reaching, preference for novel object).

**Table 2.** Functional vision evaluation.

| **Task**<br>**Domain** | **Task** | **N patients** | **Proportion** |
| --- | --- | --- | --- |
| Cognitive<br>domain | Item 3 – Pays attention to object<br>(3s) | 32 | 78.04% |
|  | Item 8 – Pays attention to object<br>(5s) | 32 | 78.04% |
|  | Item 9 – Reacts to face occlusion | 35 | 85.36% |
|  | Item 11 – Shows visual<br>preference | 28 | 68.29% |
|  | Item 12 – Habituates to object | 18 | 43.90% |
|  | Item 13 – Prefers new object | 9 | 21.95% |
|  | Item 15 – Prefers new object<br>figure | 6 | 14.63% |
|  | Item 17 – Takes object to mouth | 23 | 56.09% |
|  | Item 21 – Persistent reaching | 6 | 14.63% |
| Motor<br>domain | Item 2 – Eyes follow moving<br>person | 31 | 75.60% |
|  | Item 3 – Eyes follow plastic ring<br>(horizontal) | 22 | 53.65% |
|  | Item 4 – Eyes follow plastic ring<br>(vertical) | 23 | 56.09% |
|  | Item 7 – Eyes follow plastic ring<br>(circular) | 18 | 43.90% |
|  | Item 8 - Head follows plastic ring | 19 | 46.34% |
|  | Item 9 – Eyes follow moving<br>ball | 14 | 34.14% |

To relate functional vision evaluation to acuity, we identified which patients performed at least 2/3 of the tasks successfully. No relationship could be established between VA and the cognitive domain tasks (Acuity Regression Coefficient = −0.87 to 0.43 Log-Odds change; LR = 0.475; p = 0.4905), but there was a statistically significant relationship between VA and the motor domain tasks (Acuity Regression Coefficient = −1.00 to −0.02 Log-Odds change; LR = 4.109; p = 0.0426). Moreover, it is interesting to notice that only four children did not complete at least one task in both sets of tasks (cognitive or motor), all of them part of the most damaged group (below normative values). Most children (37) performed at least one task in the cognitive set, but 14 children did not perform even one task in the motor set. These results may imply that visual deficits in these children are impairing their motor skills even when their cognitive skills somewhat preserved. Furthermore, the children above the lower VA limit per age had a better mean performance in the cognitive (67%) and motor (66%) set than children whose VA fell below lower normative limits. Those that were marginally below the VA lower limit had showed 49% and 38% mean performance in the cognitive and motor set, respectively, while those with VA below the lower limit had 48% and 44% mean performance in the cognitive and motor set, respectively.

## 4 Discussion

The present study furthers our research team’ efforts to characterize the VA and VA development losses in children exposed to ZIKV infection during pregnancy as we did in previous studies^17,18^, examining a cohort from Jundiaí, São Paulo, Brazil’s Southeast. In this work, we evaluated a distinct population in Brazil’s Southeast, in the city of Rio de Janeiro, Rio de Janeiro.

Compared to the Jundiaí cohort, in which only few cases of microcephaly were documented, the Rio de Janeiro Cohort tells a very different story, presenting a much larger number of children with CZS in which fundoscopic alterations, VA losses and microcephaly are likely more intertwined than in the Jundiaí Cohort. At the Rio de Janeiro Cohort, only one patient did not have microcephaly (but had fundoscopic damage). 8/41 children with microcephaly did not have ophthalmologic anomalies and 7/41 children showed VA within or marginally below normative values. In other words, in this cohort, all children with VA loss also had microcephaly and/or ophthalmological damage, which makes harder to know if the VA losses in these children are due to neurologic alterations, retinal damage or both.

The children sampled for the Rio de Janeiro cohort came from a larger study, showing that beyond the fundoscopic alterations (mainly damages in the retina and optic nerve)^14^, these children also had ocular motility damage^30^, which is in agreement with the high degree of VA loss we found in this sample.

The children samples evaluated in the Rio de Janeiro Cohort and Jundiaí Cohort were close in size, with respectively 40 and 23 children in each one, but their profiles are significantly different. All subjects in Rio de Janeiro Cohort had microcephaly and/or ophthalmologic impairment against only 16 % (4/24) children with microcephaly in the Jundiaí Cohort. Most children (84%) in the Rio de Janeiro cohort had VA below or marginally below normal against 21% of children with subnormal VA in the Jundiaí Cohort. The results in our samples seem to be representative of the cohorts as a whole, given that in the Jundiaí Cohort, from 695 pregnant mothers initially accompanied, only 53 (7.6 %) were confirmed ZIKV-infected, from which only 35% of their liveborn children had confirmed ZIKV-infection and from only 4.5 % had microcephaly^39,40^.Meanwhile, in the Rio de Janeiro Cohort, from 224 infants accompanied since birth, 156 (70 %) had confirmed ZIKV-infection, from which 62 (40 %) had microcephaly^14,30,36^. In other words, despite their different sizes (Jundiaí being a much larger cohort than Rio de Janeiro), Rio de Janeiro’s children presented 10 times more confirmed ZIKV-infected children, and in the infected children a 10 times greater chance of presenting microcephaly.

Even though these differences may be partly due to different selection criteria by the IFF Fiocruz research team (while the Rio de Janeiro Cohort followed only symptomatic pregnant mothers or children with suspected ZIKV-infection, the Jundiaí Cohort followed pregnant women irrespectively of presence of ZIKV-infection symptoms), this alone does not explain the different numbers of microcephaly and fundus alteration between the two samples, nor the differences in ZIKV-infection and microcephaly incidence between them. These differences may be due to differences in virus strains^41,42^ present in the two states. They may also be due to differences of immunologic resistance between the two populations, due to previous exposure to other viral agents, nutritional profile or genetic differences^26^.

Considering that most (95%) children evaluated here were below the lower VA normative values per age, it is important to note that 26 % were only marginally below that limit. This, coupled with the fact that most children (92%) performed at least one task in the visual function sets, suggests some of them, even though heavily visually impaired, may benefit of visual rehabilitation programs, which might contribute to mitigate some of the visual damages suffered.

Only binocular evaluations were performed due to time constraints, which makes it difficult to directly relate the VA outcomes with retinal damage, since visual losses in the affected eye in monocular lesions might be compensated by the unaffected eye. Additionally measured VA varied widely in the investigated sample, and attentional, neurological and eye motility issues (such as nystagmus) may have contributed to this variability. Several precautions were taken to minimize such factors (displaying the cards vertically for children with horizontal nystagmus and testing at a 38 cm distance for all children), but those potential extraneous influences must be considered.

Most children (87 %) assessed in our study were older than 24 months at the time of testing. No studies dedicated to CZS have reported VA measurements for this age range yet. VA assessments at this age range have fewer developmental sources of variability and might offer a VA measurement that best captures visual function of those patients at school age and later in adulthood. However, since this age range has a very slight increase in VA development it was not possible to evaluate the VA development of this sample.

## Data Availability

All data produced in the present study are available upon reasonable request to the authors

